# Quantifying hope: An EU perspective of rare disease therapeutic space and market dynamics

**DOI:** 10.1101/2024.07.21.24310776

**Authors:** Emmanuelle Cacoub, Nathalie Barreto Lefebvre, Dimitrije Milunov, Manish Sarkar, Soham Saha

## Abstract

Rare diseases, affecting millions globally, pose a significant healthcare burden despite impacting a small population. While approximately 70% of all rare diseases are are genetic and often begin in childhood, diagnosis remains slow and only 5% have approved treatments. The UN emphasizes improved access to primary care for these patients and their families. Whole-genome sequencing (WGS) and next-generation sequencing (NGS) offer hope for earlier and more accurate diagnoses, potentially leading to preventative measures and targeted therapies. This study explores the therapeutic landscape for rare diseases, analyzing drugs in development and those already approved by the European Medicines Agency (EMA). We differentiate between orphan drugs with market exclusivity and repurposed existing drugs, both crucial for patients. By analyzing market size, segmentation, and publicly available data, this comprehensive study aims to pave the way for improved treatments and a brighter future for rare disease patients.

**Funding:** This work received no external funding. External agencies or companies had no role in the idea and study design, model execution and evaluation, and drafting of figures and manuscript.

## Introduction

Rare diseases pose an unprecedented disease burden to patients, doctors and caregivers impacted by a heterogeneous and conspicuous group of 7000 indications, affecting a small number of individuals (< 1 in 2000) ^1^. A recent report highlights that all rare diseases, when combined, affect one in 16 people worldwide ^2^. An estimated 300 million people are affected worldwide with a rare disease, facing neglect and marginalization, particularly in low-and middle-income countries. Roughly 80% of these diseases are genetically predisposed ^3^, with 70% showing an onset during childhood years ^4^. However, 95% of these indications lack approved treatments, diagnosis takes an average of 4.8 years and 30% of the pediatric population die before age 5 ^1^. The 2021 resolution of the United Nations (UN) on rare diseases highlights the need for improved access to healthcare, especially primary care, for the affected population and their families ^5^. Whole-genome sequencing (WGS) along with improvement in several other genetic screening techniques hold promise for presymptomatic and more accurate diagnosis of rare diseases ^6^. Next generation sequencing (NGS), non-invasive prenatal testing, carrier screening and advanced bioinformatics approaches for variant identification have been some of the key approaches applied ^7^. Non-coding DNA sequencing by WGS with these advancements can lead to earlier diagnoses, allowing for preventative measures, targeted therapies, and potentially improved outcomes ^8^. In addition, "out of hospital diagnosis tools" such as Accelrare by Sanofi and RDK by Tekkare greatly enhance the ability to diagnose patients by physicians. High sensitivity and specificity for diagnosing rare neurological disorders have been demonstrated in this retrospective diagnostic accuracy study ^9^. Integrated multi omics approaches have been shown to speed the diagnosis timeline in synergy with traditional methods ^10^. Early diagnosis is crucial for improved health outcomes and quality of life, reducing treatment costs and ease financial hardships. The limited market potential for rare disease drugs discourages pharmaceutical development. Policies like the 1983 US Orphan Drug Act ^11^ incentivize orphan drug development, leading to hundreds of approved drugs globally. Novel therapeutic strategies like gene therapy offer hope, but accessibility remains a major hurdle. Treatments like Hemgenix ^12^, Zynteglo ^13^ are exorbitantly expensive, reflecting the affordability crisis. Rare diseases often require specialized treatments available only in high-income countries, forcing some patients to pursue unaffordable medical tourism ^14^. Parents of the pediatric population and caregivers face substantial burdens, including limited information, reduced treatment options, financial strain, and potential career sacrifices ^15^. Additionally, rare diseases can lead to social stigma due to lack of awareness, visible symptoms, and inadequate support from institutions like schools and workplaces. Despite recent advancements, rare diseases continue to pose significant challenges. Prioritizing early diagnosis, effective treatment, and increased research funding through specific policies ^16^ is crucial. A multifaceted approach integrating rare diseases into healthcare systems, including mental health considerations, and raising awareness is essential for progress and meeting the unmet needs of this population.

This study explores the therapeutic interventions’ landscape against rare diseases taking into consideration 211 drugs approved by the European Medicines agency (EMA till 2021) for rare diseases or intended for rare disease use and drugs in developmental pipeline against rare diseases from 43 leading biopharmaceutical companies/biotech working in the space as shown in Table 1. We have broadly classified the EMA approved drugs for rare diseases into two segments, depending on their designation status:

(1) Drugs with Orphan Drug Designation (ODD) ^17^: These drugs received regulatory incentives and market exclusivity due to their specific development for rare diseases from EMA.
(2) Non-Orphan Drugs for Rare Diseases: Existing medications repositioned to treat rare conditions, potentially offering a faster route to patient and market access.

This expanding therapeutic landscape plays a crucial role in improving the cost of illness (COI) and quality of life (QoL) for millions of patients suffering from rare diseases in Europe. The revenue analysis encompasses the global market for these drugs. This comprehensive approach provides a broader understanding of the expanding landscape of these treatments both from a development and financial point of view. These meta analyses take into account publicly available data sources like:

(1) Drug developmental Pipelines along with financial statements of major pharmaceutical and biotechnology companies (Table 1, Table 2) and
(2) European Medicines Agency (EMA) approved drugs information until 2021 (major present pharmaceutical assets) which ensures data integrity, coverage and facilitates future research endeavors (Table 3-6).

The market share for rare disease therapeutics is segmented based on:

(1) Pharma-Specific market shares: Examining the market position of pharmaceutical companies with these therapies.
(2) Disease-Specific market shares: Understanding the market proportions of the analyzed rare diseases targeted.

These additional segmentations provide deeper insights into the market dynamics of this realm, help in better understanding the market leaders and facilitate exploration of potential developmental and market opportunities. By analyzing market segments, global revenue, and utilizing publicly available data, this study aims to concise, structure and review valuable knowledge of this evolving field.

Interventions against rare diseases approved by the EMA have a global market of around 170 billion US dollars as of the fiscal year 2022-2023. There are around 611 unique interventions that were analyzed in this study, which consists of 211 approved interventions by EMA and 400 interventions that are in developmental pipelines of 43 different pharmaceutical players. The leaders of the market are Janssen Pharmaceuticals, Abbvie, Hoffmann-La Roche AG, Bristol-Myers Squibb, Vertex Pharmaceuticals and Novartis and this estimate aligns with the 2028 estimate of the leading pharmaceutical companies worldwide by projected orphan drug sales share ^18^.

## Methods

### Data sources

Data were extracted from publicly available information on company websites on developmental pipelines. The information on finances and market were derived from public information on company financial statements. All data sources and links are summarized in Table 1.

### Meta analysis

A meta-analysis study was performed to elucidate the landscape of approved and in-development therapeutic interventions for rare diseases, taking into account the research and development (R&D) pipelines of 176 biopharmaceutical companies/biotech (Table 1). Some of the key outputs analyzed in this study include the drug name, targeted disease, the mechanism of action of the interventions, therapy areas in which these interventions fall into, collaboration status and phase of clinical development. Furthermore, the locations of the key players were analyzed to obtain a comprehensive view of the geographical space.

The second study takes into account the approved interventions in the EMA, with and without ODD status, and intended for usage in rare diseases. The annual financial reports (2023) of companies holding the market authorization of the interventions were analyzed for the product specific revenue, product specific percentage of market shares, indication specific market share distribution, company specific and disease area specific market share distribution (Table 3-7).

This multifaceted data analysis of market share and corporate pipelines delineates the rare disease therapeutic landscape and elucidates market dynamics in rare diseases.

### Identification of interactions between indications and companies

To gain a global understanding of how a company’s developmental pipelines impact the R&D landscape for a specific rare disease and the therapeutic axes, we employed a bipartite network analysis. First, we constructed a network where companies are represented as nodes and make up the first layer of the network. These company nodes were connected to two distinct sets of nodes on the other side. The first layer containing the company nodes is then connected to two additional layers. The nodes in the second layer represent company collaborators which the companies in the first layer may work with (or that were acquired as a result of mergers and acquisitions). The nodes in the third layer represent diseases currently under development by the companies the in the first layer. In situations where a pharmaceutical company, is solely responsible for both drug discovery a direct connection between the first and third layers was made. This bipartite structure allows us to capture not only the company’s research focus but also their collaborative landscape.

We then removed individual companies from the first layer of the network, simulating a company going out of business or withdrawing from a particular research area. To assess the impact of this removal, we analyzed the interactions in the resulting network, considering two key factors. Firstly, we quantified the total number of companies remaining that would actively remain in the specific disease of interest. Secondly, we analyzed the number of unique mechanisms of action (MoA) represented by the companies. By analyzing the diversity of MoA, we assessed the potential impact on the range of therapeutic approaches being explored for the disease. This combined measure offered a comprehensive understanding of how a company’s developmental pipelines affect both the overall research effort and the richness of approaches (MoA) for a particular disease (Table 8).

### Graphing and visualizations

A scatter plot visualized the relationships between various parameters, including market share, revenue of top-selling assets, competitive index, and collaborative index. The data was aggregated and collected in Microsoft Excel, using Excel functions such as *vlookup*. The plotting was done using Prism v10.2.3 (GraphPad) and custom-made scripts using Python programming language. Figure panels were prepared in affinity designer and Inkscape softwares.

## Results

### Global map of pharmaceutical players, therapy areas of interventions and their corresponding mechanisms of action (MoA)

The global distribution shows that the pharmaceutical entities involved in the development and approval of drugs for rare diseases are headquartered mostly in first world countries. The United States is the leading location for the headquarters followed by European nations, Japan and Australia (Fig 1A, B). Some of the leading countries hosting these headquarters are the Netherlands, Germany, France, Ireland, Sweden, Italy, United Kingdom, Spain, Switzerland and Belgium (Fig 1C).

**Figure 1:**
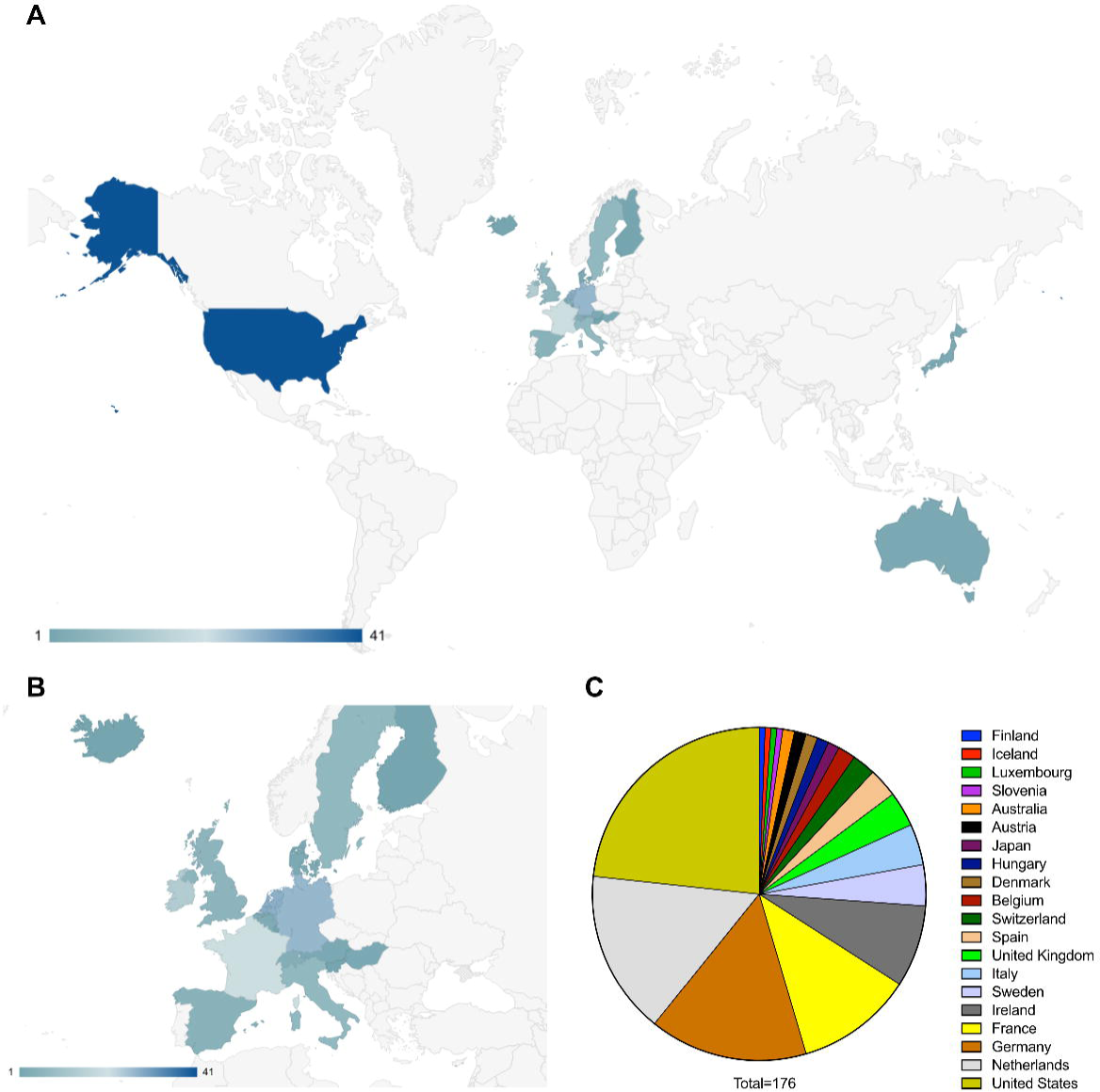
(A) Worldwide distribution of pharmaceutical entities having approved and in development therapeutics against rare diseases. (B) European distribution of pharmaceutical entities having approved and in development therapeutics against rare diseases. (C) Distribution of countries according to the number of pharmaceutical entities headquartered there.

The global leaders in therapy area distribution of the interventions against rare disease in development and approved are targeted therapy, enzyme replacement therapy (ERT), gene therapy, combinatorial therapy, antisense RNA therapy, CAR-T cell therapy and siRNA mediated interventions (Fig 2A). The mechanism of action landscape encompasses enzyme Inhibitors (such as BTK, TYK2, TTR, UBE3A-ATS, Bcl-2, VEGFR inhibitors and others), activation therapies (such as CFTR, orexin 2 receptor and GLP-2R activators), antibody-based therapies (such as anti-PD-1, anti-type I interferon receptors, anti-CD20, anti-IL-6 and anti-complement system C5), Alternative therapeutic strategies such as recombinant human clotting factor VIII, DNA alkylating agents, CAR T-cell therapy targeting B-cell maturation antigen, and drugs that modulate cellular processes like protein translation (elF2B activation) and fibrosis (TGF-β inhibition) are also implicated(Fig 2B). In the course of this study, 641 rare disease interventions, 530 unique MoAs and 25 unique modalities were analyzed, along with individual pipeline analyses for the major players in rare diseases (Supplementary Fig 1, Supplementary Fig 2). Connectivity across parent companies sponsoring the development of an intervention, their collaboration and partnering with other companies/ biotechs and the targeted disease shows that collaborations are a key component of drug development in rare disease therapies (Fig 2C). Hoffmann-La Roche AG, Takeda, Abbvie, Biogen, Affinia therapeutics, Sarepta therapeutics, PTC therapeutics, Ionis pharmaceuticals, Astrazeneca AB, Spark therapeutics, LogicBio therapeutics and Denali therapeutics take the lead in partnerships and collaborations in developmental pipelines (Fig 2D), in key therapeutic areas such as systemic lupus erythematosus (Novartis, Hoffmann-La Roche AG, Abbvie, Janssen, Astrazeneca and Bristol-Myers Squibb (including Celgene)), multiple myeloma (Ipsen Pharmaceuticals, Hoffmann-La Roche AG, Abbvie, Novartis, Janssen, Astrazeneca, Pfizer and Bristol-Myers Squibb), duchenne muscular dystrophy (Hoffmann-La Roche AG, Vertex Pharmaceuticals, Ultragenyx, Affinia therapeutics, Sarepta therapeutics, Pfizer, PTC Therapeutics and Biomarin International), amyotrophic lateral sclerosis (Biogen, Ionis Pharmaceuticals, Abbvie, PTC therapeutics, Alchemab therapeutics, Pharmanext and Denali therapeutics), IgA nephropathy (Hoffmann-La Roche AG, Alexion (part of Astrazeneca), Travere therapeutics, Astrazeneca AB, Novartis and Takeda) and spinal muscular atrophy (Hoffmann-La Roche AG, Novartis, Biogen, Ionis Pharmaceuticals and PTC therapeutics). All these players have either single or multiple programs targeting the indications. In the case of multiple programs, the corresponding pharmaceutical companies are represented by multiple occurrences (Fig 2E). The competitive landscape of drug development for rare diseases was further granulated using:

i. Single development: Only entity developing intervention against specific indications;
ii. Competitors: Multiple entities involved in therapeutic interventions development against the indication;
iii. Unique mechanism of action (MoA): Number of unique therapeutic axes used for therapeutic development from a mechanistic point of view and
iv. Shared MoAs: Number of shared therapeutic axes employed for therapeutic development from a mechanistic point of view.

**Figure 2:**
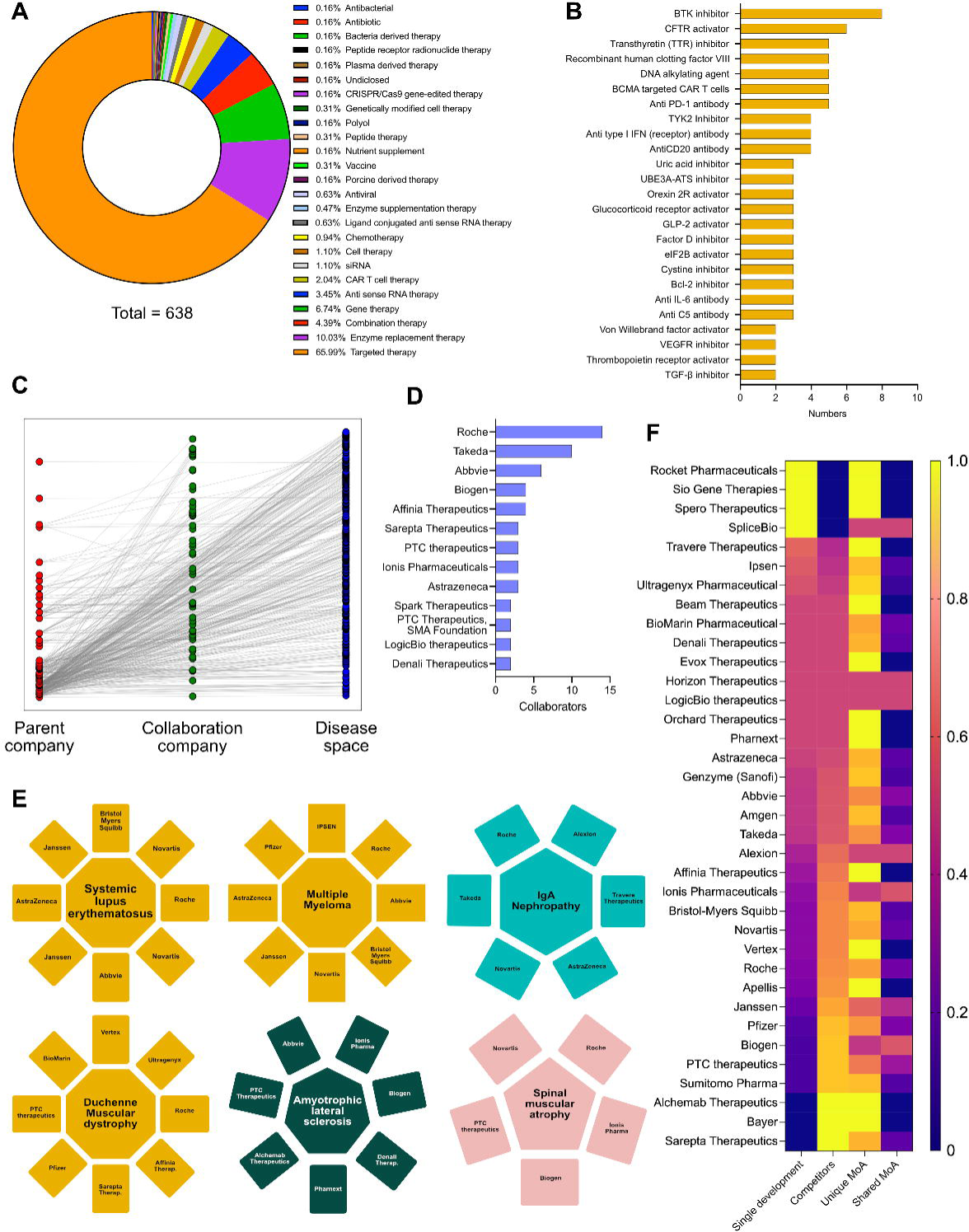
(A) Intervention specific therapy area distribution in the therapeutic landscape. (B) Intervention specific mechanism of action (MoA) distribution of the therapeutic landscape. (C) Connectivity matrix connecting the parent company, collaborative companies and the diseases targeted in their developmental pipeline. (D) Top pharmaceutical companies having the highest number of collaborations in their developmental pipelines. (E) Top 6 disease areas dominated by multiple pharmaceutical players increasing competitiveness. (F) Heatmap showing the innovative index and competitive index (disease and MoA) of different pharmaceutical players having developmental pipelines in rare disease space (Yellow: Highest; Blue: Lowest).

We were able to conclude that Rocket pharmaceuticals, Sio Gene therapies, Sphero therapeutics and SpliceBio have a high degree of innovative index, but very low degree of competitive index. Companies like Sarepta Therapeutics, Bayer AG, Alchemy Therapeutics, Sumitomo Pharmaceuticals, Janssen, Biogen, Hoffmann-La Roche AG, PTC therapeutics, Vertex Pharmaceuticals, Ionis Pharmaceuticals, Affinia Therapeutics, Alexion (part of Astrazeneca AB) have a low degree of innovative index and a very high degree of competitive index, while Abbvie, Amgen, Genzyme (part of Sanofi), Astrazeneca AB, Orchard therapeutics, LogicBio Therapeutics have a comparable innovative and competitive index (Fig 2F). Furthermore, analyses of all the companies involved in therapeutic development in unique axes of MoAs yields biotech entities with high degree of innovative index or competitive index (either from disease perspective or MoA perspective) and are potentially key determinants for M&A activities (Fig 2F).

### Global distribution of EMA approved drugs (until 2021) with and without ODD status against rare indications having market shares

The EMA has approved interventions against 113 indications with ODD and 201 indications without ODD status till 2021. 13 of these indications (multiple myeloma, acute lymphoblastic leukemia, type 1 Gaucher disease, pulmonary arterial hypertension, idiopathic pulmonary fibrosis, hereditary angioedema, adrenal insufficiency, chronic lymphocytic leukemia, follicular lymphoma, nephropathic cystinosis, Fabry disease, cystic fibrosis and haemophilia B) have known interventions, with both ODD and non-ODD status (Fig 3A). Out of the 400 interventions approved against rare diseases by EMA, 35% have ODD status while 65% do not have ODD status (Fig 3B). There has been a continuous increase in the number of drug approvals against rare disease with and without ODD status in the last few decades (Figure 3C, 3D). The EMA approved drugs having ODD against rare diseases are dominated by targeted therapy (71.5%) and enzyme replacement therapy (8.5%) (Fig 3E). Some of the key indications targeted by ODD drugs are multiple myeloma, cystic fibrosis, spinal muscular atrophy, endogenous cushing syndrome, hereditary angioedema and acute myeloid leukemia (Fig 3F). Novartis, Biomarin International Limited, Janssen Cilag International, Hoffmann-La Roche AG, Celgene (acquired by Bristol-Myers Squibb), Pfizer, Alexion (Astrazeneca partner), Alnylam pharmaceuticals, Chiesi Pharmaceutical, Incyte Biosciences, Shire (part of Takeda), Takeda pharmaceuticals, Vertex pharmaceuticals, Advanced Accelerator Applications,Akcea Therapeutics, Amgen, Bayer AG, Bluebird bio, Genzyme (part of Sanofi) and Ipsen Pharmaceuticals are the key pharmaceutical players in this group (Fig 3G).

**Figure 3:**
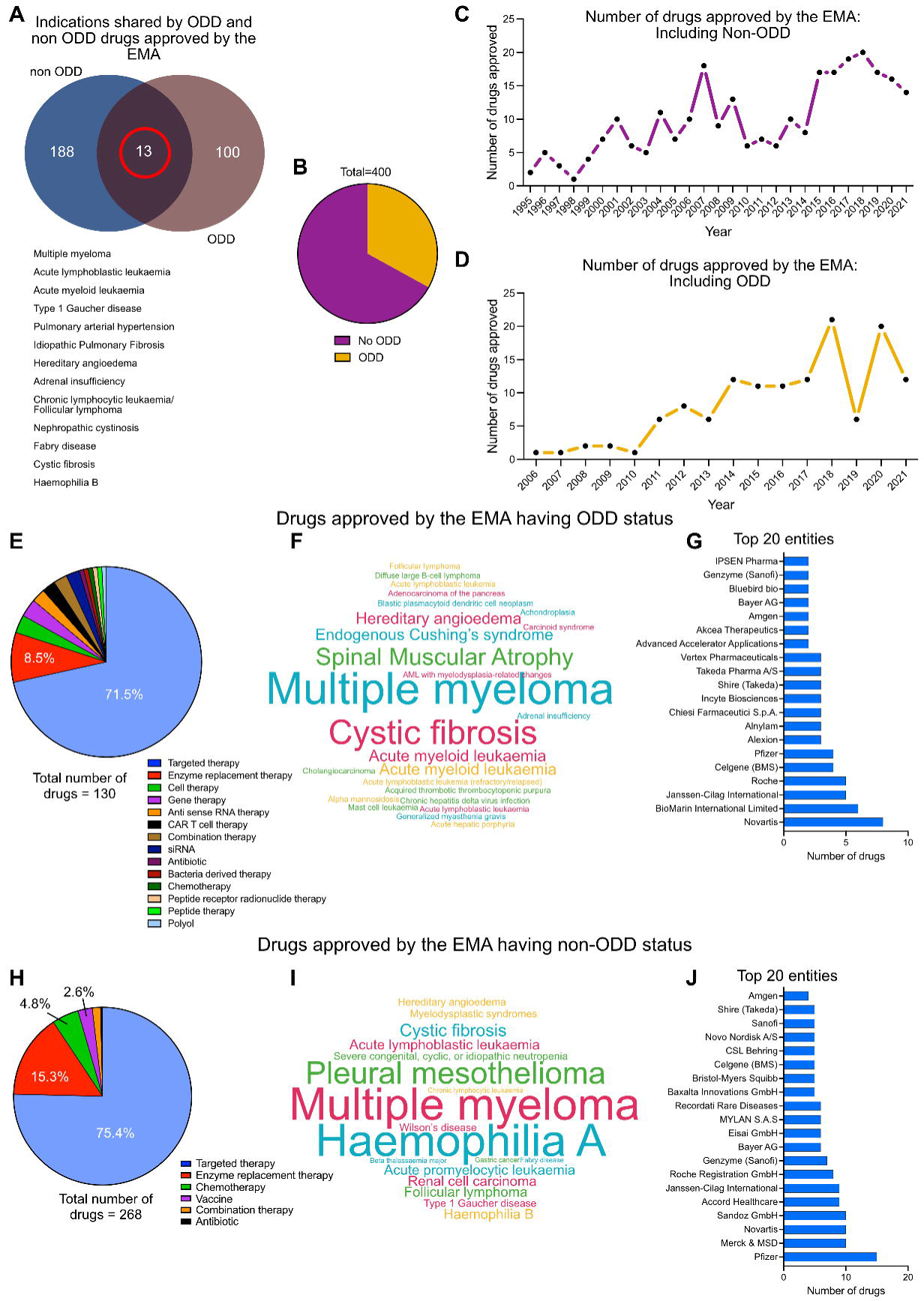
(A) Indications shared by EMA approved drugs for rare diseases with and without ODD. (B) Distribution of total number of EMA approved drugs having ODD and no ODD. (C) Approval timeline of drugs against rare diseases by EMA without ODD (1995-2021). (D) Approval timeline of drugs against rare diseases by EMA with ODD (2005-2021). (E) Therapy area distribution of drugs with ODD (n=130). (F) Cloud diagram showing the distribution of rare indications against which the ODD designated drugs were being approved (G) Top 20 pharmaceutical entities involved in this intervention space with ODD. (H) Therapy area distribution of drugs without ODD (n=268). (I) Cloud diagram showing the distribution of rare indications against which the non-ODD drugs were being approved (J) Top 20 pharmaceutical entities involved in this intervention space without ODD.

The EMA approved drugs against rare diseases without ODD status are dominated by targeted therapy (75.4%), enzyme replacement therapy (15.3%) and chemotherapy (4.8%) (Fig 3H). Some of the key indications targeted by non-ODD drugs are multiple myeloma, haemophilia A, pleural mesothelioma, cystic fibrosis and acute lymphoblastic leukemia (Fig 3I). Pfizer, Merck, Novartis, Sandoz, Accord healthcare, Janssen International, Hoffmann-La Roche AG, Genzyme (part of Sanofi), Bayer AG, Eisai Gmbh, Mylan SAS, Recordati Rare Diseases, Baxalta Innovations, Bristol Myers Squibb, Celgene(acquired by Bristol-Myers Squibb), CSL Behring, Novo Nordisk, Sanofi, Shire (part of Takeda) and Amgen remain key players in the non-ODD group (Fig 3J).

### Chemical network and mechanism of action driven landscape analysis of approved drugs against rare indications with and without ODD designation

Drug-drug(red lines) and drug protein (green lines) interaction networks represent the chemical landscape of the interventions. It shows that the drugs are quite sparse spatially which is due to the presence of innovative interventions targeting different biomarkers in the rare disease space which has received ODD (Fig 4A). Some of the key mechanisms of action represented by the ODD designated drugs are small molecule targeted therapy (such as CFTR activator, transthyretin (TTR) inhibitor, cysteine inhibitor, FXR inhibitor, TGF-β inhibitor, 16S/23S rRNA inhibitor, 30S ribosomal protein S12 inhibitor, 5-HT2 receptor agonist, σ1 receptor inhibitor, ALAS1 inhibitor), antibody-mediated targeted therapy (such as anti CD19 antibody, anti IL6 antibody, anti C5 antibody, anti CCR4 antibody, anti CD19/CD3 bispecific antibody, anti FGF23 antibody, anti CD20 antibody, anti GD2 antibody, anti-PA (*B. anthracis* toxin) antibody, anti-plasma kallikrein antibody, anti von Willebrand factor (vWF) antibody), antibody-drug conjugate (such as anti CD30 antibody/monomethyl auristatin E (MMAE), anti CD22 antibody/ Calicheamicin, anti CD33 antibody/ Calicheamicin) and gene therapy (such as AAV mediated SERPINA1 gene transfer, AAV9 mediated SMN gene transfer, AAV mediated RPE65 gene transfer) (Fig 4B).

**Figure 4:**
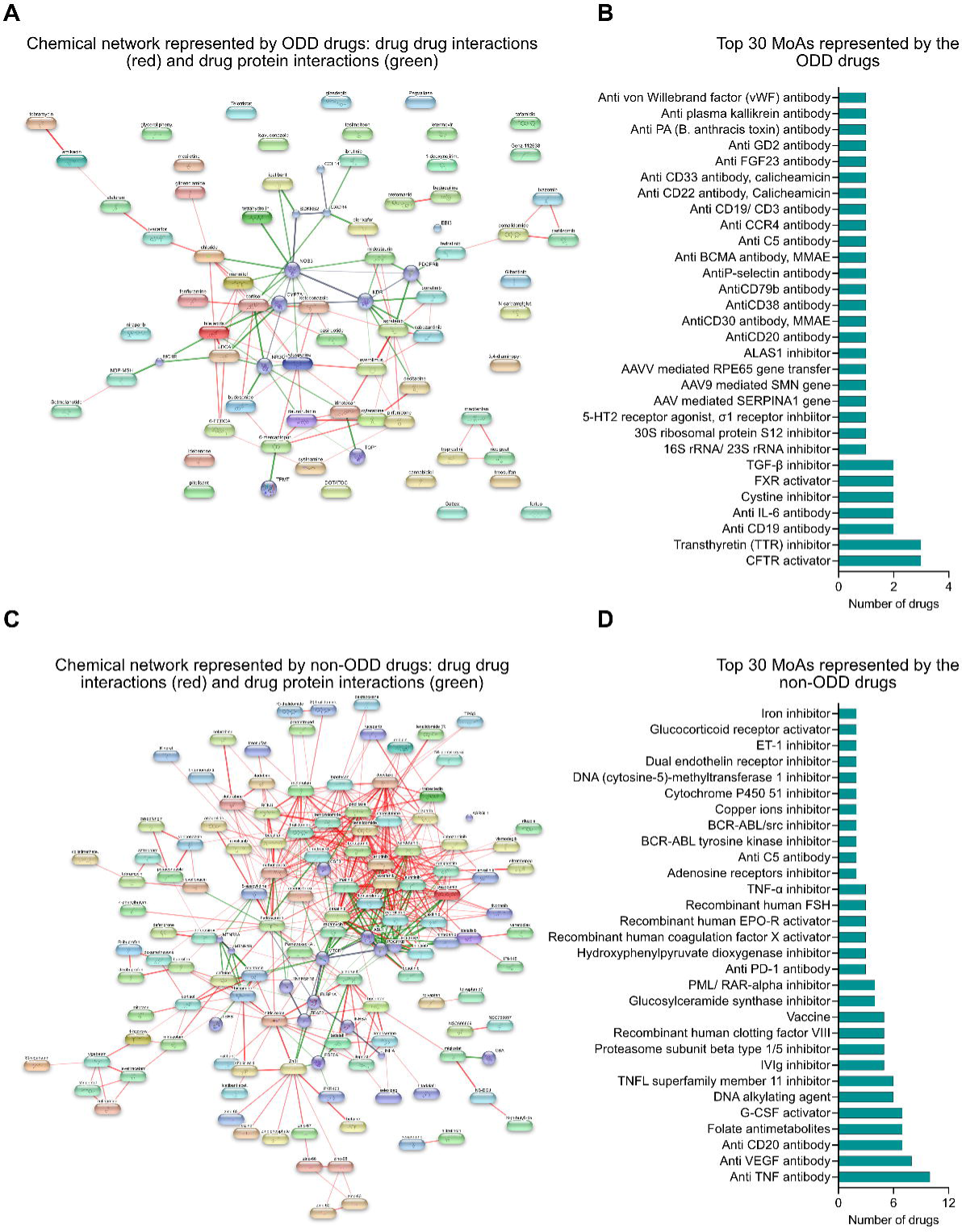
(A) Chemical network represented by ODD designated interventions: drug-drug (red) and drug-protein (green) interaction framework. (B) Top 30 mechanisms of actions represented by interventions with ODD designation. (C) Chemical network represented by non-ODD interventions: drug-drug (red) and drug-protein (green) interaction framework. (D) Top 30 mechanisms of actions represented by interventions without ODD designation.

Similarly, drug-drug(red lines) and drug-protein (green lines) interaction networks among the non-ODD interventions might suggest that these interventions are mostly repositioned from other non-rare indications that share common targets and mechanisms of action (Fig 4C). The key mechanisms of action represented by the non-ODD designated drugs are small molecule targeted therapy, antibody mediated targeted therapy, enzyme replacement therapy and alternative therapies like DNA alkylating agent and folate antimetabolites (Fig 4D).

### Market share landscape of therapeutic space against rare diseases with and without ODD status

A review of the pharmaceutical industry identified 20 key players by revenue and market share specific to rare diseases. Janssen (10.8%) came first in the list, followed by Hoffmann-La Roche (9.6%) and AbbVie (9.1%). Mergers and acquisitions are reflected, with Celgene (6.8%) included under Bristol-Myers Squibb and Alexion (4.2%) under AstraZeneca. The remaining companies include Vertex Pharmaceuticals (5.5%), Novartis (6.5%), Amgen (4.4%), Pfizer (4.4%), Merck (4.3%), CSL Behring (3.3%), Pharmaxis (2.4%), Genzyme (part of Sanofi, 2.2%), Boehringer Ingelheim (2.1%), Takeda (2.1%), Biogen (2.0%), BioMarin (1.3%), Eisai (1.1%), and Eli Lilly (1.0%) (Figure 5A, 5B). In addition, Amgen, Novartis, Hoffmann-La Roche AG, Pfizer, Biomarin international, Celgene (part of Bristol-Myers Squibb) Genzyme (part of Sanofi), Janssen, Merck, Recordati Rare Diseases, Sandoz, Takeda, Bristols-Myers Squibb, Alexion (part of Astrazeneca), Bayer AG, Biogen, CSL Behring, Eli Lilly, Vertex Pharmaceuticals Abbvie and Alnylam Pharmaceuticals also boast a large pharmaceutical portfolio of assets (Fig 5C). The market leaders of this therapeutic space were estimated using a correlation study between the total revenue and the highest grossing asset of each of the companies. It showed that there are three separate divisions among the players: The **market leaders** (6-12%): Janssen, Abbvie, Hoffmann-La Roche AG and Novartis and Celgene (part of Bristol-Myers Squibb); **The upcoming major players** (2.5-6%): Vertex pharmaceuticals, Bristol-Myers Squibb non-Celgene portfolio, Pfizer, Amgen, Merck, Alexion (part of Astrazeneca), CSL Behring and Astrazeneca AB non-Alexion portfolio; **The challengers** to the market space (<2.5%): Pharmaxis, Boehringer Ingelheim, Takeda, Biogen, Genzyme (part of Sanofi), Bayer AG and Alnylam Pharmaceuticals among others (Fig 5D). Physiological system specific company distribution of assets were analyzed and it provides the granularity of the number of interventions in correlation to the body system their targeted indications affect. A further analysis by organ system unveiled a diverse distribution of interventions. Several companies, like AbbVie, Amgen, Bristol-Myers Squibb, Janssen, Novartis, Pfizer, Roche, and Sandoz focus their efforts with a single intervention specific to the complex liver/ heart/ lung or the broader reproduction/ lung/ stomach/ gut/ skin systems. Similarly, companies like Amgen, Bristol-Myers Squibb, Janssen, and Sanofi hold a single intervention in the kidney/ blood/ gut/ bone space, suggesting a more targeted approach in these areas.

**Figure 5:**
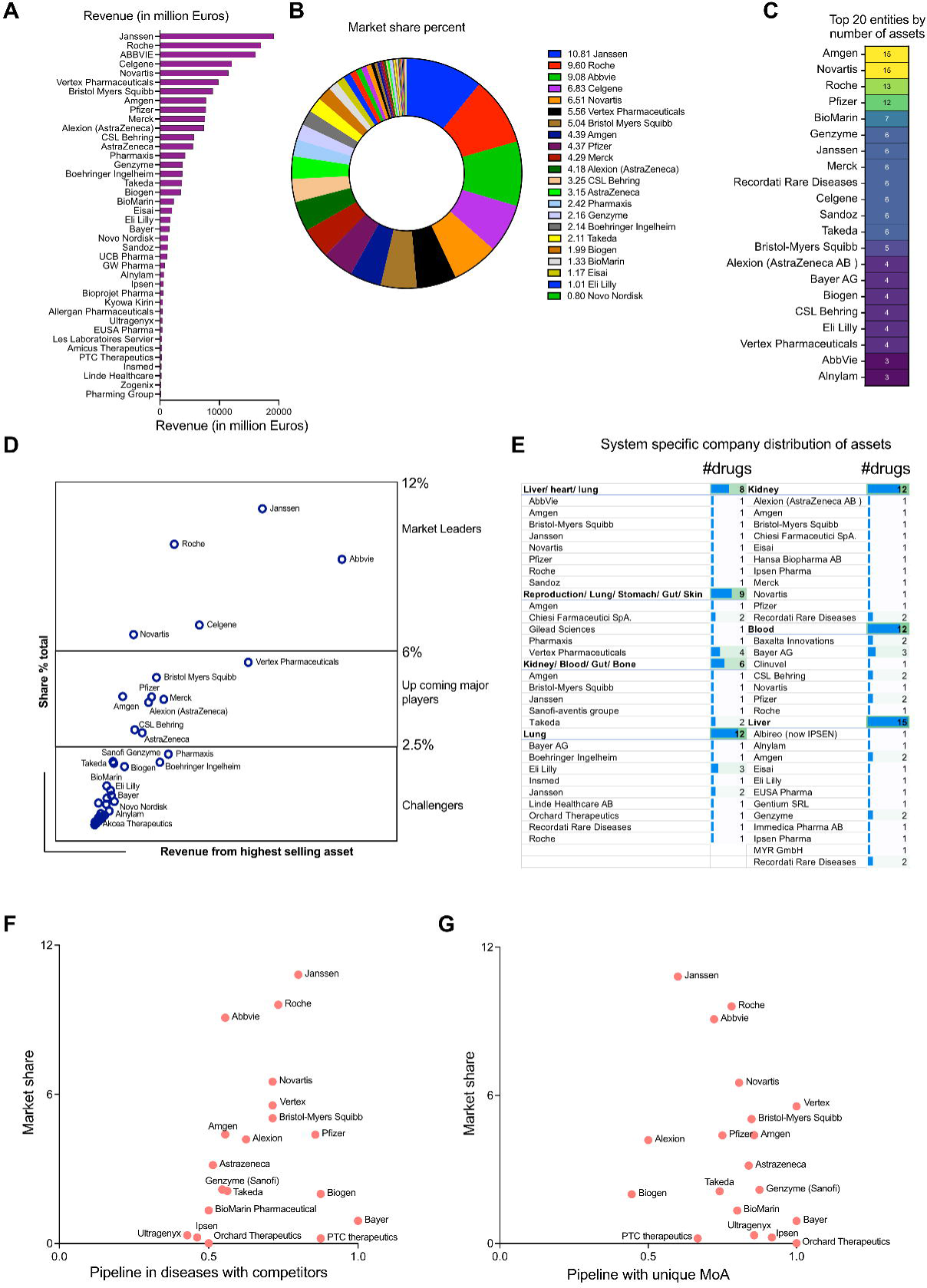
(A) Revenue distribution among the pharmaceutical entities (in million dollars) in rare disease space. (B) Rare disease therapeutic market share distribution among the pharmaceutical entities (percentage). (C) Top 20 pharmaceutical companies having the highest number of money making assets (D) Market leader estimation using the corresponding correlation coefficients between the total revenue and highest grossing asset. (E) Physiological system specific company distribution of assets. (F) Scatter plot showing the competitive index for disease space targeted by the pharmaceutical players (developmental pipelines) in perspective to the total market shares (approved drugs). (G) Scatter plot showing the innovative index driven by unique MoAs targeted by the pharmaceutical players (developmental pipelines) in perspective to the total market shares (approved drugs).

However, the landscape shifts when examining companies with multiple interventions within specific organ systems. Here, we see a more concentrated focus. For example, Janssen and Eli Lilly take a leading role in the lung system with two and three interventions, respectively. Similarly, Bayer AG, Recordati Rare Diseases, and Amgen (among others) hold multiple assets in the blood and liver systems, indicating a strategic investment in these critical areas. This trend highlights the variety of approaches companies carry out when developing interventions, with some opting for a broader reach and others focusing on specific organ systems with multiple offerings (Fig 5E).

We defined the competitive index of a company based on quantitative assessment of the existing competitors in the therapeutic space. If a therapeutic strategy had more than one player, it was deemed as competitive. On the other hand, if the mechanism of action of the therapeutic asset was unique to a particular company, it could indicate high innovation index. A scatter plot between the total market share of the pharmaceutical entities in rare therapeutics space and their total assets in pipeline shared by other competitors gives an estimate of the market leaders in two perspectives: Approved drugs in market and drug development in a competitive environment Fig 5D). Janssen, Hoffmann-La Roche AG and Novartis are the leaders having the highest market shares and richest developmental pipeline in the competitive disease environment (Competitive index closer to 1). Market leaders like Abbvie have a high share of the market space but play in a lower risk environment (Competitive index close to 0.5). The upcoming players like Vertex Pharmaceuticals, Bristol Myers Squibb (including Celgene developmental portfolio), Pfizer and Alexion (part of Astrazeneca AB) have median market shares and also are a part of a highly competitive disease landscape (Competitive index closer to 1). Amgen and Astrazeneca (without Alexion portfolio) have median market percentages but target diseases which are shared to a lesser extent among the competitors (Competitive index closer to 0.5). Biogen, Bayer AG and PTC therapeutics are in a strong competitive position (Competitive index closer to 1), but have comparatively less market shares than its competitors. Entities like Genzyme (part of Sanofi), Takeda, Biomarin Pharmaceuticals, Ipsen Pharmaceuticals, Ultragenyx and Orchard therapeutics are working in therapeutic areas which can accommodate higher levels of competition (Competitive index closer to 0.5) and despite their market shares being lower, they have the opportunity to expand their market positioning (Fig 5F). Analyzing the unique mechanism of action of the interventions in the developmental pipeline targeting rare diseases, all the prominent market players like Vertex Pharmaceuticals, Hoffmann-La Roche AG, Abbvie, Takeda Novartis, Bristol-Myers Squibb (including Celgene portfolio), Amgen, Pfizer, Astrazeneca AB, Genzyme, Biomarin International have a very high innovation index (close to 1) and are distributed across the entire market share landscape (high to low). Entities like Janssen, Alexion (part of Astrazeneca AB), Biogen, PTC Therapeutics cover are distributed in the market share landscape having relatively lower innovation index (close to 0.5) (Fig 5G).

### Market share landscape of disease and system specific therapeutics against rare diseases

An analysis of disease specific market share was performed taking into account the total revenues of multiple drugs against the indications. We implemented a bubble diagram to visualize the relationships which shows the top indications with the highest market share given their approved therapies: cystic fibrosis, multiple myeloma, polyarticular juvenile idiopathic arthritis, renal cell carcinoma, haemophilia A, spinal muscular atrophy and chronic lymphocytic leukemia (Fig 6A) A physiological system (single or multi system) specific market share analysis takes into account the total cash flow from interventions targeting the diseases affecting the specific systems which was represented by a radial map (Scale: Blue: Highest market share; Brown: Lowest market share) (Fig 6B). The top organ systems having the highest pharmaceutical assets are in multiple organs encompassing the liver, heart, lung, stomach, gut, skin and others such as the kidney, blood, bone, musculoskeletal systems, or affecting distinct systems such as reproduction, liver and heart, among others (Fig 6C).

**Figure 6:**
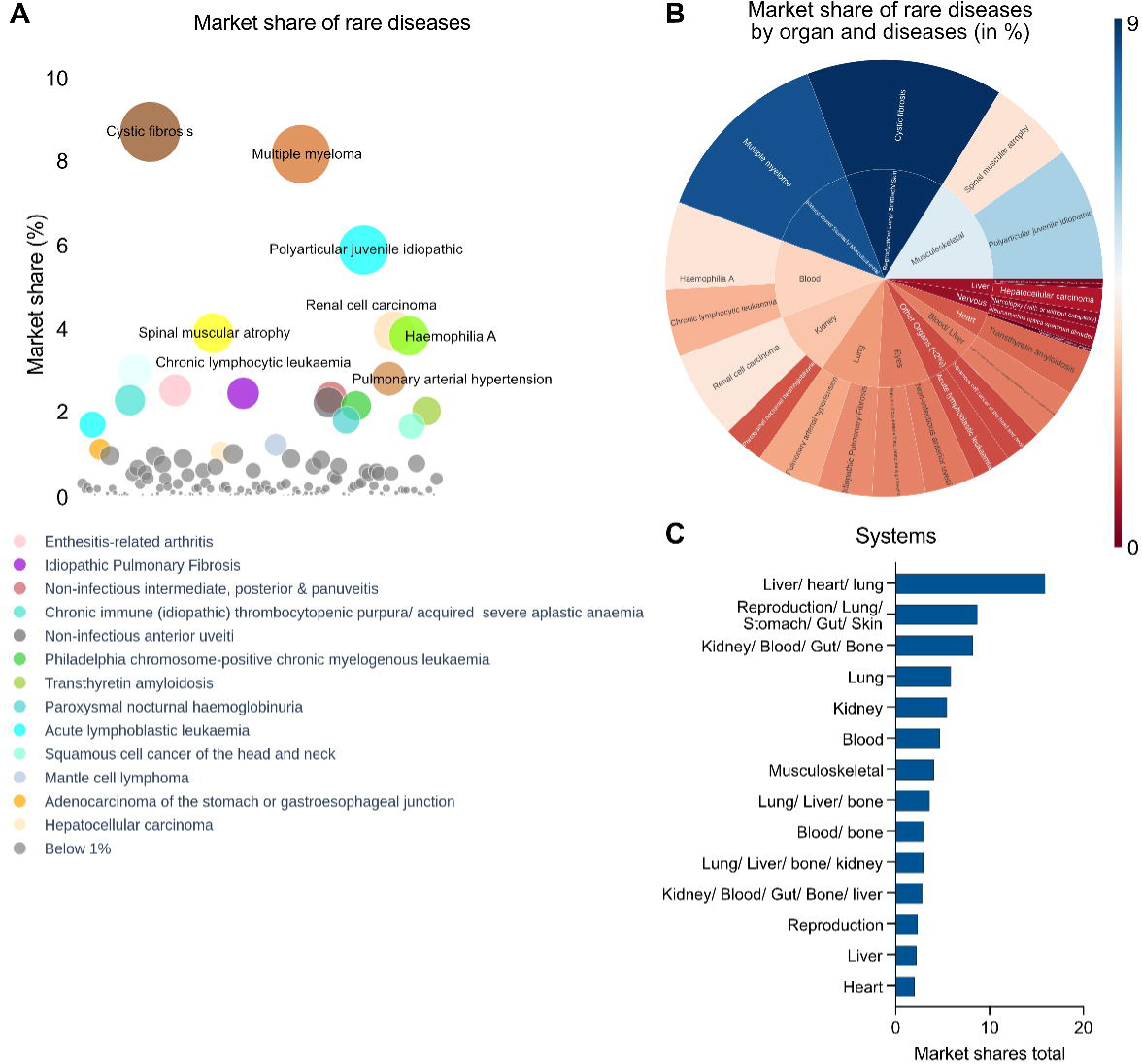
(A) Disease specific market share analysis of approved interventions (EMA) in rare disease therapeutics space. (B) Radial map showing disease and organ specific market share analysis (Blue: Highest; Brown: Lowest) of approved rare disease therapeutics in the EMA. (C) Top disease specific organ systems targeted by EMA approved drugs in context of market shares.

## Discussion

For those living with a rare disease, the world can feel like a vast, uncharted territory. The challenges are immense-limited treatment options, a lack of awareness, and the constant struggle to find answers. However, recent research offers a promising light. Global efforts to combat these complex conditions are gaining momentum, revealing a dynamic field driven by innovation. The study reveals a concerted international effort, where pharmaceutical giants in the United States, Europe, Japan, and Australia are leading the charge, leveraging their expertise and resources to develop life-changing therapies. This international collaboration is crucial, as rare diseases often transcend geographical boundaries. By pooling resources and sharing knowledge, researchers are accelerating the pace of discovery, bringing much-needed hope to patients worldwide. The research delves deeper, uncovering the specific therapeutic strategies employed to combat this condition. Targeted therapies, as demonstrated by the existing drug pipelines across biotechs and pharma, are emerging as the most dominant approach. These therapies are exquisitely precise, honing in the specific biological pathways that underlie rare diseases ^19^. Unlike traditional medications with broader applications, targeted therapies offer a more potent and well-tolerated approach. This focus on precision medicine represents a significant leap forward in the battle against rare diseases ^20^. While targeted therapies stand as the leading force, the treatments against rare disease doesn’t stop there. Enzyme replacement therapies ^21^, for instance, offer a lifeline for patients with conditions caused by missing or dysfunctional enzymes. Similar to a missing cog in a complex machine – enzyme replacement therapies act like replacement parts, restoring functionality and alleviating symptoms. Gene therapy also holds immense promise in targeting specific genetic predispositions responsible for the indications ^22^. This modality tackles the root cause of some rare diseases by introducing healthy copies of defective genes directly into a patient’s cells. While still in its early stages, gene therapy has the potential to offer permanent cures for a range of rare conditions. As previously mentioned, the therapeutic landscape further differentiates between drugs with and without ODD ^17^. ODD status for drugs is intended at incentivizing research and development in this underserved field. The analysis reveals a fascinating distinction between these two categories. ODD drugs form a more distinct network of interactions, suggesting they target unique biological pathways not addressed by existing medications. This highlights the cutting-edge nature of ODD drugs – they represent the frontiers of rare disease research, to discover groundbreaking best-in-class or first-in-class therapies. The concept of a drug-protein network provides a compelling way to visualize the intricate interactions between medications and the human body. The stronger the interaction between a drug and a protein, the thicker the connecting line. In the case of ODD drugs, these networks appear more discrete, showcasing their unique targeting mechanisms while for non-ODD drugs, the network is denser and more connected, reflecting the life cycle management (LCM) and drug repositioning opportunities ^23^. While scientific innovation holds the key to unlocking new treatment options, the commercial landscape plays a critical role in ensuring these therapies reach the patients who need them most. This study delves deeper into the market share percentages, revealing the pharmaceutical giants who are leading the generated revenue. Companies like Janssen, Roche, AbbVie, Bristol-Myers Squibb and Novartis are at the forefront, demonstrating their commitment to rare disease research and development. However, the landscape is not static. Emerging players like Amgen and Vertex Pharmaceuticals are making significant strides, bringing fresh perspectives and innovative approaches to the table. This healthy competition is a positive sign for patients, as it fuels the development of even more effective therapies. The increasing number of players also suggests a growing awareness of the importance of this field, paving the way for a brighter future for patients with rare diseases. One exciting possibility lies in precision medicine ^24^, where targeted therapies are becoming even more granular ^25^. Researchers envision tailoring therapies to the unique genetic fingerprint of each patient or patient group ^26^. This personalized approach promises to maximize efficacy, minimize side effects, and usher in a new era of individualized treatment. Additionally, gene therapy advancements like CRISPR ^27–29^, AAV-mediated therapies ^30–32^, and antisense oligonucleotide approaches ^33,34^ offer the potential for permanent cures through precise genetic code editing. Finally, the vast amount of data generated by research presents a unique opportunity ^35^. By harnessing the power of big data analytics, researchers can identify hidden patterns and connections, accelerating drug discovery and our understanding of rare diseases ^36^. While the future looks promising, there are still hurdles to overcome. One of the biggest challenges is ensuring equitable access to these life-changing therapies ^37^. The high cost of rare disease medications can be a significant barrier for patients ^38,39^. Collaborative efforts between researchers, pharmaceutical companies, and policymakers are crucial to bridge this gap and ensure that everyone has the opportunity to benefit from these advancements. The landscape of rare disease research is an ecosystem filled with innovation and collaboration. Research into diverse range of therapeutic approaches, from targeted therapies to gene therapy, are constantly pushing the boundaries of science. In the face of a healthcare crisis for millions, philanthropic funding stands as a power of progress for those living with rare diseases. Unlike traditional pharmaceutical companies whose focus is on maximizing profits, philanthropic organizations like rare disease dedicated foundations ^40–42^ and private philanthropists ^43^ tries to bridge the critical gap between the available therapies and the small market size associated with rare diseases. This allows for crucial early-stage research that might otherwise be left unexplored, potentially leading to groundbreaking discoveries. Philanthropic activities have guided development of the state of the art care against multiple cancer types in the USA led by healthcare centers like Dana Farber Cancer research Institute, MD Anderson comprehensive cancer center and Sidney Kimmel comprehensive cancer center ^44–46^. These hospitals and cancer centers today, being some of the best healthcare centers worldwide for oncologic indications, all started in the last century when philanthropy paved the way to increased research, large scale patient care and access. It is thought that rare disease should follow a similar trajectory. Furthermore, philanthropic funders are driven by a mission of alleviating human suffering, not just financial gain. This patient-centric approach allows them to champion research on diseases affecting smaller populations, a demographic often overlooked by for-profit entities. This commitment to patient well-being extends beyond just funding. Philanthropic organizations can also have a vital role in fostering innovation by supporting smaller research institutions and companies with promising ideas that might not fit the traditional model. By providing crucial financial resources and fostering collaboration, philanthropic funding acts as a catalyst, accelerating the development pipeline and bringing life-changing treatments closer to the millions living with the daily challenges of rare diseases. There have been recent advancements in the EU pharmaceutical reforms ^16^ where Members of the European Parliament (MEPS) have adopted proposals for sweeping reforms to EU pharmaceutical legislation, aiming to stimulate innovation while improving drug accessibility and affordability. Rare and chronic diseases are the key areas of focus. The European Union (EU) has proposed a comprehensive strategy to address the challenges of rare diseases. This initiative ensures the development and availability of high-quality, safe, and effective treatments for all EU patients with rare diseases. Equitable access to these treatments across all member states is a key focus, aiming to remove geographical barriers. Additionally, the proposal emphasizes the importance of a secure supply chain to guarantee uninterrupted patient access to these potentially life-saving therapies. Finally, fostering a robust research environment is envisioned to stimulate the continuous development of novel treatments for this complex group of diseases. The proposal aims to target some of the important shortcomings in the present rare pharmaceutical legislation:

(1) patient unmet medical needs, including those of children, with rare diseases are consistent.
(2) health systems face significant challenges in coping up with the exorbitant price-tags of the medications.
(3) patient access to rare disease medications through the EU is incoherent.
(4) regulatory hurdles serve as a roadblock in many cases leading to increased delay in patient access.

The European Union is aiming to simplify its regulations for all medicines by merging specific rules for rare disease and pediatric treatments into a single, more coherent framework. Importantly, this change won’t affect the high quality, safety, and effectiveness standards for these specialized medicines as they will still follow the same rigorous approval process as any other medication. However, the new regulations will recognize the unique challenges of developing treatments for rare diseases and children. Unlike medications for common illnesses, market forces alone don’t create enough incentive for research in these areas. Therefore, the revised regulations will maintain specific requirements that provide additional support for this crucial research. This ensures continued development of new and effective treatments for these often-overlooked patient populations. To ensure the best outcome of revising rare disease and children’s medicine regulations, a comprehensive analysis was conducted. This included examining three different policy directions (A, B, and C) for each regulation. They are as follows: Option A: Maintains 10-year exclusivity and adds a transferable voucher for high unmet medical need (HUMN) products. This voucher grants a 1-year extension or can be sold for use on another product. Option B: Eliminates the current 10-year exclusivity for all rare disease treatments.

Option C: Offers variable exclusivity (5, 9 or 10 years) based on the type of treatment (established use, new drug and HUMN respectively). An additional one-year extension is possible for HUMN and new drug products based on patient access across the EU. Following a comprehensive evaluation of the specific objectives and potential economic and social impacts, Option C emerged as the preferred policy choice. It strikes a balance between all four revision goals, promoting investment and innovation, especially for high unmet medical need (HUMN) treatments. This won’t hinder the development of other rare disease drugs. Additionally, Option C is expected to (i) boost EU pharmaceutical competitiveness, including small and medium-sized enterprises (SMEs), (ii) improve patient access to medicines through earlier entry for generic and biosimilar drugs and (iii) reduce administrative burdens and better accommodate new technologies with more flexible criteria for defining orphan diseases. The proposed revisions target core procedures required for market authorization of medicines and their life cycle management activities resulting in a streamlined regulatory framework with improved efficiency, effectiveness and access ^47^.

Patients associations are crucial for raising awareness against rare diseases ^48,49^. Rare diseases for obvious reasons are not well known by the common people due to their meager incidence rates. Patient associations can take up initiatives to educate people leading to higher awareness among people and increased focus among healthcare experts as well as policymakers for rare disease treatments. Thus they play a vital role in driving the remission of the high unmet needs that are associated with rare diseases. In addition to that, patients associations contribute substantially to funding and research against rare diseases driven by non-profit organizations, biotech SMEs and big pharmaceutical entities ^50^. Diving more into details, clinical trials play a major role in therapeutic development against all types of indications. However, is it challenging to meet the eligibility criteria for clinical trials targeting rare diseases given the difficulty associated to patient identification and enrollment, as there is a low number of volunteers for orphan drug development. Patients association helps considerably in solving this issue by connecting clinical researchers working on clinical trials to specific and marginalized patient populations. This facilitates successful development of the intervention and expanded patient access. Successful collaborations between patient associations and clinical researchers (CT sponsors) can help in elucidating the high unmet needs of patients suffering from rare diseases and their caregivers. This type of partnership would help in optimize the experience of patients participating in targeted clinical trials which has the potential to improving patient quality of life (QoL) significantly by improvement of the standard of care treatments ^51,52^.

### Limitations of this study

This study has two potential limitations. First, as it is a EU perspective of the rare disease therapeutics landscape, it includes only the EMA approved drugs and not the FDA approvals. However, the expanded intervention landscape is a future scope of study. Second, some approved interventions have multifaceted disease targets, both rare and non-rare. This might have led to some overlapping market shares from non-rare diseases being incorporated in the total market size estimation for the EMA approved rare disease therapeutics.

## Data Availability

All data are available in the main text or the supplementary materials.

## Acknowledgements

We thank Dr. Pierre-Axel Monternier and Mr. Victor Hannothiaux for reading the manuscript and providing critical feedback.

## Author Contributions

Conceptualization: SS, MS

Methodology: MS, EC

Investigation: MS, EC

Visualization: NBL, DM

Supervision: SS

Writing—original draft: SS, MS, EC

Writing—review & editing: SS, MS, EC, NBL, DM

## Data and materials availability

All data are available in the main text or the supplementary materials.

## Tables

Table 1: List of data sources used in the report.

Table 2: Distribution of pharmaceutical players and biotech across the world. Table 3: List of drugs and interventions analyzed in the present study.

Table 4: List of drugs in development by companies and segregated by indications and MoA. Table 5: Landscape of rare disease therapeutics having orphan drug designation.

Table 6: Landscape of rare disease therapeutics having non-orphan drug designation. Table 7: Revenue and market share landscape for the interventions analyzed.

Table 8: List of developmental pipelines across pharma companies with the largest market shares in rare diseases.

## Supplementary materials

Supplementary Fig 1

Pipeline of assets in leading pharmaceutical companies such as Amgen, Novartis, Roche, Pfizer and IPSEN.

Supplementary Fig 2

Pipeline of assets in leading pharmaceutical companies such as Biogen, AstraZeneca/ Alexion, AbbVie, Takeda and Janssen.

