## Supplementary figures and images for "Quantifying hope: An EU perspective of rare disease therapeutic space and market dynamics"

### Supplementary fig 1

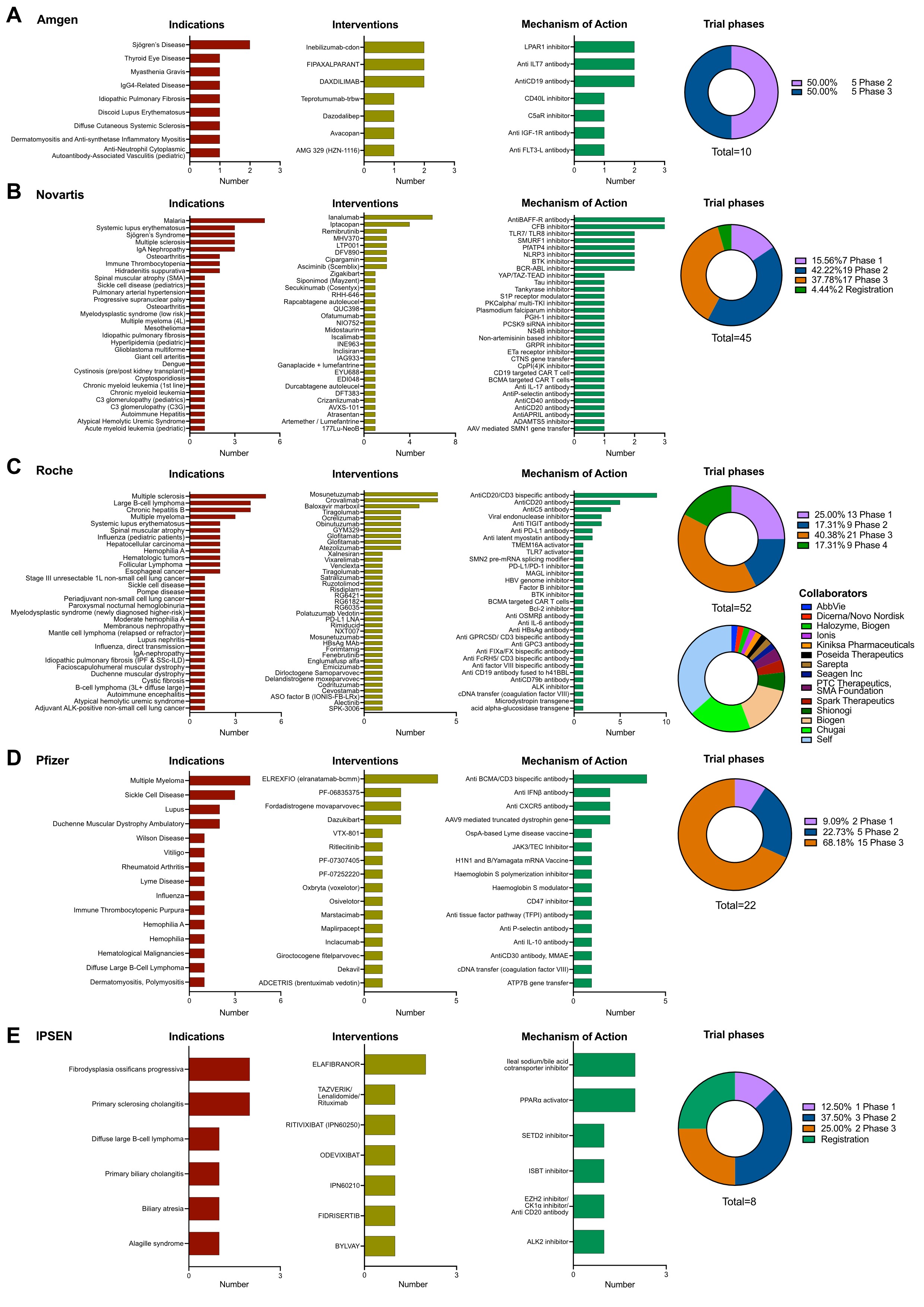

### Supplementary fig 2

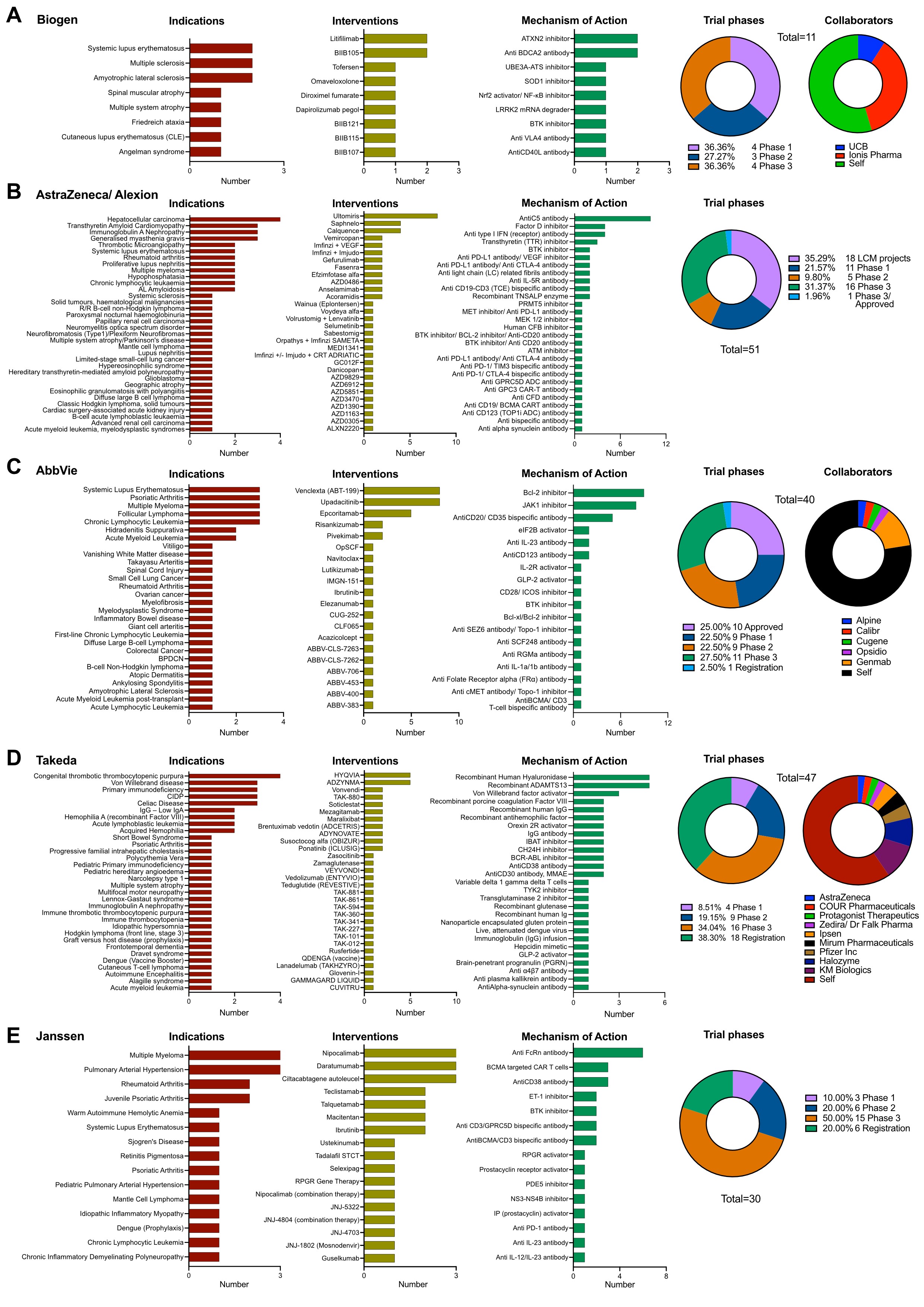
